# Expanding the Clinical and Molecular Spectrum of *TUBB2B* Through Distinct Variants Identified Across Multiple Families

**DOI:** 10.64898/2025.12.28.25342917

**Authors:** Shaghayegh T. Beheshti, Angad Jolly, Ahmed K. Saad, Haowei Du, Lauren E. Westerfield, Chloe Munderloh, Divya Kalra, Yifan Wu, Yi Chen, Marie-Claude Gingras, Shalini N. Jhangiani, Sarenur Yilmaz, Maha S. Zaki, Daniel G. Calame, Davut Pehlivan, Richard A. Gibbs, Richard A. Lewis, James R. Lupski, Jennifer E. Posey

## Abstract

*TUBB2B* encodes a β-tubulin isotype essential for neuronal proliferation, migration, and organization during brain development. Pathogenic heterozygous variants in *TUBB2B* have been associated with a range of neurodevelopmental disorders, with phenotypes including polymicrogyria and corpus callosum abnormalities. However, the phenotypic spectrum remains heterogeneous, likely influenced by variant-specific effects on microtubule formation and stability, an area that warrants further investigation. Homozygous *TUBB2B* variants are exceedingly rare, with one family reported to date.

We describe five individuals in four families with rare *TUBB2B* variants. Four variant alleles are described, including a previously reported *de novo* missense variant p.(G98R), with potential phenotypic expansion including panhypopituitarism, a previously reported *de novo* missense variant, p.(I202T) demonstrating phenotypic heterogeneity between individuals, a *de novo* missense variant, p.(Q15K) at a polyamination site critical for microtubule stability, and a novel homozygous missense variant located within a region of absence of heterozygosity in two siblings from consanguineous parents p.(V49I). Both individuals also have a pathogenic homozygous truncating variant in *ALKBH8* p.(R559Afs*56), representing a rare dual molecular diagnosis driving clinical features reflective of contributions from both genes. These cases expand the known clinical spectrum of *TUBB2B*-related tubulinopathies, illustrate intragenic phenotypic heterogeneity, including with recurrent variants and provide novel insights into potential mechanisms of disease, such as effects at polyamination sites and rare recessive inheritance, underscoring the need for nuanced genotype-phenotype interpretation in both diagnostic and counseling contexts.

## Introduction

Mammalian brain development relies on tightly regulated neuronal proliferation, migration, and differentiation, all supported by microtubules (MTs), which drive mitosis, cell migration, and intracellular transport^1^. MTs are built from αβ-tubulin heterodimers that assemble into polar protofilaments, oriented with α-tubulin at the minus end and β-tubulin at the plus end^2^. MT dynamics and interactions with MT-associated proteins (MAPs), including motor proteins and regulators of MT stability, are vital for neurodevelopment. The tubulin GTP-binding pocket is key for folding, dimer stability, and nucleation. Disruptions in these structural or functional elements can result in a group of neurodevelopmental disorders collectively known as tubulinopathies^3^.

Pathogenic variants in 12 of 19 human α-, β-, and γ- tubulin genes (*TUBA1A, TUBA4A, TUBA8, TUBB1, TUBB2A, TUBB2B, TUBB3, TUBB5, TUBB4A, TUBB8*, *TUBGCP2*, and *TUBG1*) are linked to rare disorders, affecting the central nervous system^2,4^. These often involve neuronal migration defects, agenesis of the corpus callosum, structural abnormalities of the basal ganglia, and hindbrain malformations^2^. Notably, even within a single gene, different variants can lead to distinct phenotypes due to their variable effects on MT function^5,6^. Variant alleles causing these pathogenic proteins can be functionally categorized as: (I) reduced supply of functional tubulin heterodimers, (II) altering GTP binding, (III) modifying protofilament interactions, or (IV) disrupting MT interactions with motor proteins or MAPs^7^.

*TUBB2B*, expressed predominantly in the brain, exemplifies this functional and phenotypic diversity. *TUBB2B* is predicted to be loss-of-function (LOF) intolerant (pLI = 1). Heterozygous missense variants in *TUBB2B* are linked to cortical dysplasia, complex with other brain malformations (CDCBM7 [MIM:610031]) with variable molecular mechanisms of disease such as LOF, dominant negative, and gain of function. This diversity is further reflected in the phenotypic heterogeneity seen with the same *TUBB2B* variant, as illustrated by a family carrying the c.530A>T (p.Asp177Val) mutation: the healthy carrier parent only showed childhood language delay, while the affected son had mild neurodevelopmental delay and tubulinopathy-consistent MRI findings, and a fetus was severely affected with microcephaly and major brain anomalies^7^. The only reported homozygous *TUBB2B* p.(R390Q) variant was found in three individuals with Uner Tan syndrome from a consanguineous Turkish family^8^. Functional studies showed preserved heterodimer formation but reduced MT stability, highlighting the varied functional impacts of *TUBB2B* variants^8^.

Here, we report four unrelated families with *TUBB2B* variants. Three families have heterozygous missense variants: one previously reported p.(G98R) and now associated with panhypopituitarism, demonstrating a phenotypic expansion, another with the recurrent p.(I202T) variant, located at a functionally predicted critical position and illustrating the phenotypic heterogeneity that can arise even from the same variant, and another at a key polyamination site, p.(Q15K), important for microtubule stability^9^. Additionally, we describe a consanguineous family previously reported with a homozygous truncating variant in *ALKBH8* p.(R559Afs*56)^10^, for whom a novel homozygous missense variant, *TUBB2B* p.(V49I), is also now identified in the affected siblings. We present detailed clinical and molecular findings, further expanding the genotypic and phenotypic spectrum of *TUBB2B*-related disorders (Table 1).

**Table 1.** Clinical features of individuals with *TUBB2B* variants annotated using Human Phenotype Ontology (HPO) terms. Table 1 summarizes phenotypic findings in five affected individuals across four families carrying rare missense variants in the *TUBB2B* gene. Phenotypes are categorized by clinical system and annotated using Human Phenotype Ontology (HPO) identifiers. Growth parameters are reported as standard deviation (SD) scores where available. Yes indicates that the phenotype was documented in the clinical record. No indicates the feature was absent and blank cells indicate the feature was not reported. Cases 4A and 4B are siblings with a homozygous variant in *TUBB2B*, while Cases 1, 2, and 3 each harbor *de novo* heterozygous variants. ABR - Auditory brainstem response; EMG - Electromyography; NCV - Nerve conduction velocity; NH₃ - Blood ammonia; AA - Plasma amino acids; OA - Urine organic acids; Tandem MS - Tandem mass spectrometry; MR spectroscopy - Magnetic resonance spectroscopy; ECHO - Echocardiogram; FISH - Fluorescence in situ hybridization; SNRPN - Small nuclear ribonucleoprotein polypeptide.

| Phenotype Category | Phenotype | HPO ID | Case 1 | Case 2 | Case 3 | Case 4A | Case 4B |
| --- | --- | --- | --- | --- | --- | --- | --- |
| Demographics | Age range |  | 1-5 year | 5-10 year | 1-5 year | 1-5 year | 10-15 year |
|  | Sex |  | Male | Male | Male | Male | Female |
|  | Pregnancy and Delivery |  | Full term, no complications | Full term, no complications | Full term, no complications | Full term, no complications | Full term, no complications |
| Neurological Findings | Corpus Callosum Atrophy | HP:0007371 | Yes (Agenesis) | Yes (Hypogenesis) | Yes (Agenesis) | Yes (Agenesis) | Yes (Agenesis) |
|  | Ventriculomegaly | HP:0002119 |  | Yes |  | Yes | Yes |
|  | Delayed CNS myelination | HP:0002188 |  |  |  | Yes | Yes |
|  | Cerebral Hypoplasia | HP:0006872 | Yes |  |  |  |  |
|  | Cerebellar Hypoplasia | HP:0001321 |  | Yes |  | Yes | Yes |
|  | Cortical Dysplasia | HP:0002539 | No | Yes | No | No | No |
|  |  |  |  | (Abnormal gyration, Thick cortex) |  |  |  |
|  | Thinning of brain stem |  |  | Yes (mainly pons) |  |  |  |
| Seizures/Epilepsy | Seizure | HP:0001250 | Yes | No | Yes | No | No |
| Developmental | Intellectual Disability | HP:0001249 |  | Yes |  | Yes | Yes |
|  | Motor delay | HP:0001270 | Yes | Yes | Yes | Yes | Yes |
|  | Speech delay | HP:0000750 | Yes | Yes | Yes | Yes | Yes |
|  | Global developmental delay | HP:0001263 |  | Yes |  | Yes | Yes |
| Behavioral Features | Autism | HP:0000717 |  |  | Yes | Yes |  |
|  | Happy demeanor | HP:0040082 |  |  | Yes |  |  |
|  | Attention deficit hyperactivity disorder | HP:0007018 |  |  |  | Yes |  |
|  | Mood swings |  |  | Yes |  |  |  |
|  | Hyperactivity | HP:0000752 |  | Yes |  |  | Yes |
| Vision/Ophthalmological Findings | Optic nerve hypoplasia | HP:0000609 | Yes |  |  |  |  |
|  | Astigmatism | HP:0000483 | Yes |  |  |  |  |
|  | Nystagmus | HP:0000639 | Yes |  |  |  |  |
| Craniofacial Findings | Dysmorphic facial features | HP:0001999 |  | Yes | Yes | Yes | Yes |
| Other Neurological Features | Microcephaly | HP:0000252 | Yes | Yes |  |  |  |
|  | Cerebral palsy | HP:0100021 | Yes |  |  |  |  |
|  | Hypotonia | HP:0001252 |  | Yes | Yes | Yes | Yes |
|  | Hyperreflexia | HP:0001347 |  |  |  | Yes | Yes |
| Genitourinary Findings | Bilateral cryptorchidism | HP:0000028 |  |  |  | Yes |  |
| Skin Findings | Hair hypopigmentation | HP:0005599 |  |  | Yes |  |  |
| Other Medical Features | Panhypopituitarism | HP:0000871 | Yes |  |  |  |  |
|  | Delayed puberty | HP:0000823 |  |  |  |  | Yes |
| Studies performed |  |  | Prior clinical lab testing positive for the *TUBB2B* variant | Normal karyotyping, metabolic workup, fundus examination, ABR, EMG, and NCV. | Blood NH<sub>3</sub>, plasma AA, urine OA, tandem MS, and MR spectroscopy were negative. Hearing, ECHO, and EMG were normal. Karyotype was 46,XY. Angelman testing (FISH for SNRPN, methylation) was negative. |  |  |
| *TUBB2B* variant |  |  | c.292G>A: p.(Gly98Arg) | c.605T>C: p.(Ile202Thr) | c.43C>T: p.(Gln15Lys) | c.145G>A: p.(Val49Ile) | c.145G>A: p.(Val49Ile) |

## Materials and Methods

All families provided written, informed consent for publication as well as consent for participation in genomic research as appropriate^11^. Genomic DNA was extracted from blood, and trio exome sequencing was performed using established protocols^12^. Case 2 exome sequencing and variant identification was performed in a commercial laboratory. Exome capture for cases 1,3, and 4 was conducted with the Baylor College of Medicine (BCM) Human Genome Sequencing Center design (52-Mb, Roche NimbleGen, RRID: nif-0000-31466), and sequencing was performed on the Illumina HiSeq platform (∼100× coverage). Reads were aligned to the hg19 reference using the Mercury pipeline^13^, and variants were called with ATLAS2 (Case 3) or xATLAS^14^ (Cases 1 and 4) and annotated as previously described^15^.

Using the tool BafCalculator, we detected absence of heterozygosity (AOH) using unphased exome sequencing data, and used AOH as a surrogate for runs of homozygosity (ROH) (https://github.com/BCM-Lupskilab/BafCalculator)^16^. B-allele frequency was derived by comparing the proportion of alternate to total read counts at variant sites. These values were then analyzed using the circular binary segmentation algorithm^17^ to define AOH regions.

Exome data were analyzed for rare, potentially damaging variants under autosomal and X-linked (dominant and recessive) inheritance models. Variants with minor allele frequency <0.1% in gnomAD v2/v4 were prioritized, followed by *in silico* pathogenicity predictions, including Combined Annotation Dependent Depletion (CADD)^18^, AlphaMissense^19^, PrimateAI^15^, and REVEL^20^ scores. Copy number variant (CNV) analysis was performed using two exome-based CNV detection tools: XHMM^21^ and HMZDelFinder^22^.

Target regions were PCR-amplified (HotStar Taq, QIAGEN) with gene-specific primers (Table S1) and sequenced by Sanger (BCM Sequencing Core). Data were analyzed and aligned using SnapGene (GSL Biotech).

## Results

In total, five individuals from four families are described.

### Case 1

The proband is a male child in the 1-5 year age range, born following a full-term, uncomplicated pregnancy and delivery. Birth growth parameters were within the expected range. Early postnatal head circumference was between 2 to 3 standard deviation below the mean. Family history includes a relative with melanoma and another relative who was stillborn.

A head ultrasound in early infancy demonstrated agenesis of the corpus callosum and ventriculomegaly, which were confirmed by CT head and brain MRI. Optic nerve sizes were markedly reduced relative to expected, and microcephaly was raised as a potential etiology for this observation. During early childhood panhypopituitarism with central hypothyroidism and adrenal insufficiency was diagnosed. The combination of agenesis of the corpus callosum, panhypopituitarism, and optic nerve hypoplasia led to a diagnosis of Septo-Optic Dysplasia (SOD), despite the confounding observation of microcephaly. The proband additionally demonstrated global developmental delay requiring physical and speech therapy, cortical visual impairment, astigmatism, acquired nystagmus, and seizures.

Anthropometric measurements obtained in early childhood demonstrated weight and height within the expected range and head circumference more than 3 standard deviations below the mean. Brain MRI showed corpus callosum agenesis and cerebral atrophy, consistent with *TUBB2B*-related tubulinopathies. Exam revealed hypotonia and poor visual tracking. Chromosomal microarray was normal.

Research reanalysis of clinical exome sequencing demonstrated a *de novo* heterozygous missense variant in *TUBB2B* [Chr6:3226031C>T (hg19); NM_178012.5; c.292G>A, p.(G98R)], absent from gnomAD v4 and internal databases, This variant allele was predicted deleterious by multiple *in silico* models (CADD=25.3; REVEL=0.836; AlphaMissense=0.99; PrimateAI=0.94) (Table 2), and mapped to a highly evolutionarily constrained region (Figure 1B). Sanger sequencing confirmed *de novo* status of the variant in the proband (Figure 1A).

**Figure 1.**
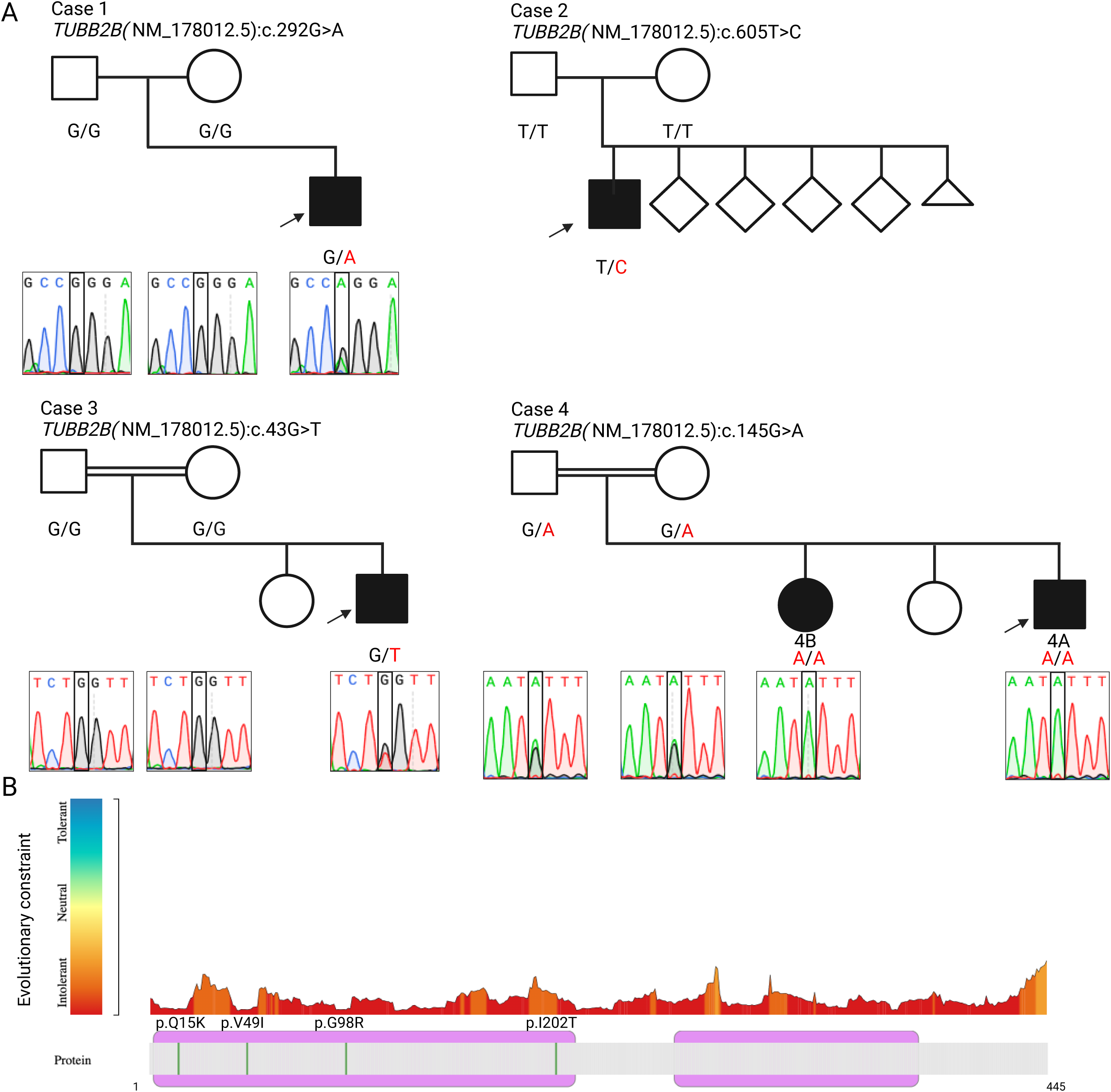
Identification and characterization of *TUBB2B* variants in four families with affected individuals. **(A)** Pedigrees for all families and Sanger sequencing chromatograms for three families with rare *TUBB2B* missense variants. Case 1 carries a *de novo* heterozygous c.292G>A p.(Gly98Arg) variant. Case 2 carries a *de novo* heterozygous c.605T>C p.(Ile202Thr) variant. Case 3 carries a *de novo* heterozygous c.43G>T p.(Gln15Lys) variant. Case 4 includes two affected siblings (Case 4A and Case 4B) with a homozygous c.145G>A p.(Val49Ile) variant, inherited from heterozygous carrier parents. Sanger sequencing confirmed the variants and their segregation within each family. **(B)** Intolerance landscape of the *TUBB2B* protein generated using MetaDome v1.0.1. Regions of the protein are color-coded based on evolutionary constraint, ranging from tolerant (blue) to intolerant (red). The four variants identified in this study p.(Gln15Lys), p.(Val49Ile), p.(Gly98Arg), and p.(Ile202Thr) are indicated by vertical green lines and are located in highly intolerant regions of the protein, supporting their likely pathogenicity. *Created with BioRender.com*.

**Table 2.** Zygosity and predicted pathogenicity scores for missense variants identified in affected individuals. This table summarizes the zygosity and *in silico* pathogenicity predictions for four missense variants in *TUBB2B* (transcript NM_178012.5) identified in five affected individuals and previously identified variant in *ALKBH8* (transcript NM_138775.3) in two affected individuals. Scores include CADD (v1.6), where values above 20 suggest deleteriousness; REVEL and AlphaMissense, both of which range from 0 to 1 with higher scores indicating greater pathogenic potential; and PrimateAI, where values above 0.8 are considered likely pathogenic. All variants are absent from the gnomAD version 4 database.

| Case ID | Gene | Transcript | Variant<br>(cDNA:protein) | Zygosity | CADD | REVEL | AlphaMissense | PrimateAI | gnomAD<br>v4 |
| --- | --- | --- | --- | --- | --- | --- | --- | --- | --- |
| Case 1 | *TUBB2B* | NM_178012.5 | c.292G>A<br>p.(Gly98Arg) | Heterozygous<br>(*de novo*) | 25.3 | 0.836 | 0.99 | 0.94 | 0 |
| Case 2 | *TUBB2B* | NM_178012.5 | c.605T>C<br>p.(Ile202Thr) | Heterozygous<br>(*de novo*) | 24.7 | 0.96 | 0.96 | 0.66 | 0 |
| Case 3 | *TUBB2B* | NM_178012.5 | c.43C>T<br>p.(Gln15Lys) | Heterozygous<br>(*de novo*) | 28.5 | 0.589 | 0.99 | 0.82 | 0 |
| Case 4A<br>Case 4B | *TUBB2B* | NM_178012.5 | c.145G>A<br>p.(Val49Ile) | Homozygous | 25 | 0.52 | 0.57 | 0.79 | 0 |
| Case 4A<br>Case 4B | *ALKBH8* | NM_138775.3 | c.1675delC<br>p.R559Afs*56 | Homozygous | - | - | - | - | 0 |

### Case 2

The proband is a male in the 5-10 year age range, born to healthy parents following an uneventful pregnancy and delivery. Birth weight and length were within the expected range, while head circumference was between 1 and 2 standard deviations below the mean. He has healthy siblings with one reported miscarriage, and no similarly affected relatives (Figure 1A).

Clinical evaluation demonstrated global developmental delay. Gross motor milestones were markedly delayed, with assisted ambulation achieved in childhood and limited independent ambulation thereafter. Expressive language was impacted, with minimal spoken words. He understood simple commands and could identify some body parts but had not achieved sphincter control. Behavioral features included hyperactivity and mood swings. There was no history of seizures. He has received physiotherapy, occupational therapy, and speech therapy since early childhood.

At examination, weight and height were within the expected range, while head circumference was more than 3 standard deviations below the mean. Dysmorphic features included a long face, narrow forehead, arched eyebrows, bilateral ptosis, puffiness of the eyelids, an upturned nose, a short philtrum, bow-shaped lips with broad, prominent upper incisors, and large, low-set ears with prominent lobules.

Neurologic assessment revealed drooling, hypotonia, and preserved deep tendon reflexes. Brain CT showed abnormal gyration, a thickened cortex, mildly enlarged lateral ventricles, hypogenesis of the corpus callosum, a small cerebellar vermis, and thinning of the brainstem, most pronounced at the pons.

Clinical exome analysis following a negative diagnostic evaluation (Table 1) identified a *de novo* heterozygous missense variant in *TUBB2B* [Chr6:3225718T>C (hg19); NM_178012.5; c.605T>C, p.(I202T)]. This variant was absent in gnomAD v4 and it affects a highly conserved residue (Figure 1B). *In silico* tools predicted a deleterious effect (CADD=24.7; REVEL=0.96; AlphaMissense=0.96; PrimateAI=0.66) (Table 2). Segregation analysis in the parents confirmed the *de novo* status of the variant in the proband (Figure 1A).

### Case 3

The proband is a male in 1-5 year age range, born to consanguineous parents following an uneventful pregnancy and delivery. Birth growth parameters were within the expected range. He has an unaffected sibling, and family history was unremarkable. (Figure 1A).

Clinical evaluation showed global developmental delay, with delayed independent ambulation. The proband had autistic features, a happy demeanor, and seizures. EEG revealed bilateral occipitotemporal abnormalities with spike wave discharges. Brain MRI showed diffuse corpus callosum hypoplasia, consistent with *TUBB2B*-related tubulinopathies. The proband also had mild dysmorphic features, including downslanting palpebral fissures, a bulbous tip of the nose, tented upper lip, and white-yellow hair depigmentation.

Research exome analysis following a negative diagnostic evaluation (Table 1) identified a *de novo* heterozygous missense variant in *TUBB2B* [Chr6:3227735G>T (hg19); NM_178012.5; c.43C>T, p.(Q15K)]. This variant was absent in gnomAD v4 and the internal BHCMG database (∼13,000 exomes) (Table 2). It affects a highly conserved residue within a known polyamination site important for microtubule stability (Figure 1B). *In silico* tools predicted a deleterious effect (CADD=28.5; REVEL=0.589; AlphaMissense=0.99; PrimateAI=0.82). Sanger sequencing confirmed the *de novo* status of the variant in the proband (Figure 1A).

### Case 4

Previously described in Saad *et al*.^10^, the proband is a male child in the 1–5 year age range, born following an uncomplicated pregnancy and delivery. He has an affected sibling and an unaffected sibling. No additional relatives were reported to have similar neurological features (Figure 1A).

Components of the proband’s phenotype which were consistent with his homozygous *ALKBH8* pathogenic variant p.(R559Afs*56), included global developmental delay, attention and speech deficits, hypotonia, and distinctive craniofacial features (Table 1). He also had bilateral undescended testes and an IQ of 51 (Stanford-Binet). Growth parameters in early childhood were between 2 and 3 standard deviations below the mean. He also had hyporeflexia and brain MRI findings not previously linked to *ALKBH8*, including mild ventriculomegaly, volume loss, cerebellar hypoplasia, corpus callosum thinning, and abnormal myelination (Figure 2B).

**Figure 2.**
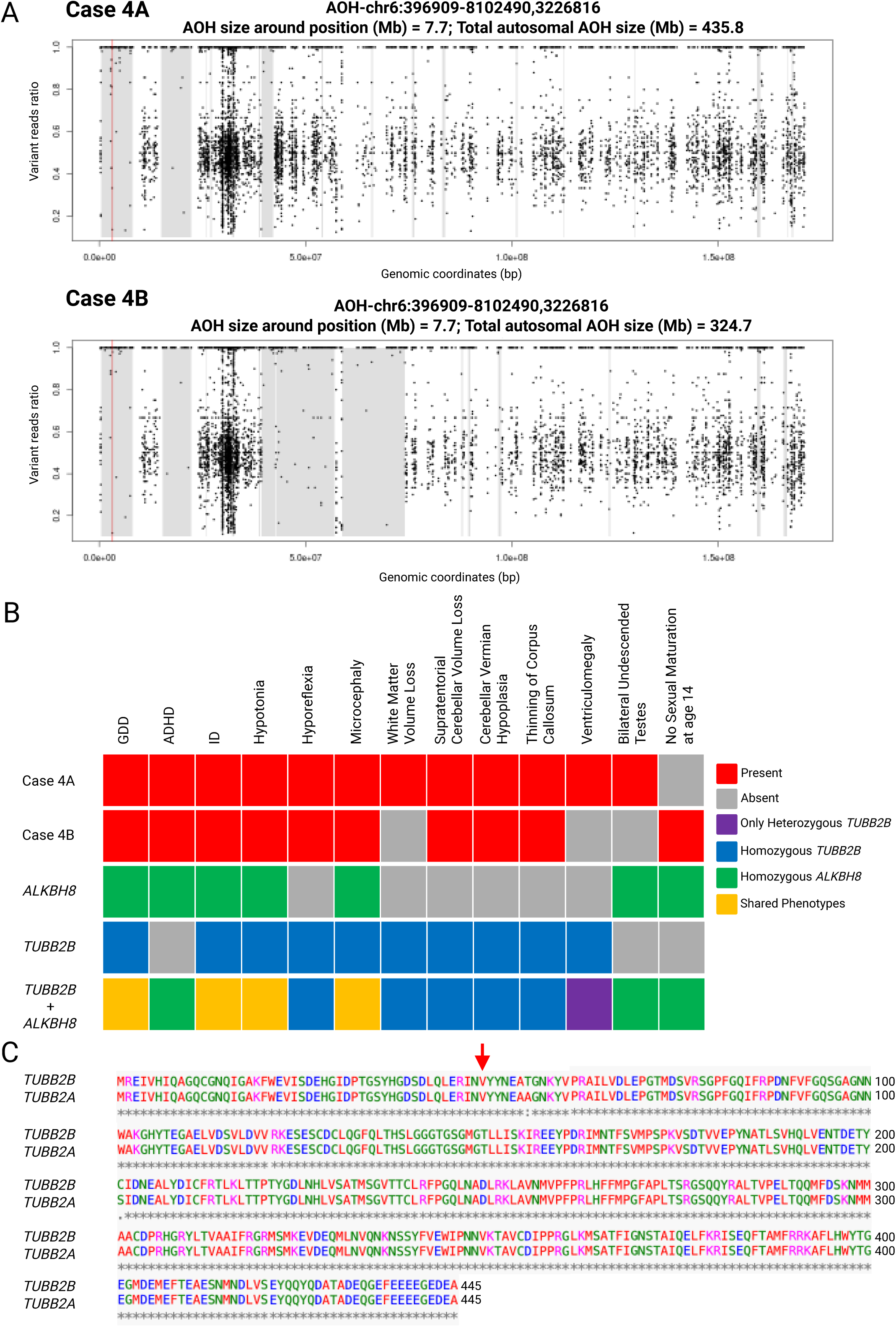
AOH plots and Genomic and phenotypic evidence supporting dual molecular diagnosis in siblings with homozygous *TUBB2B* and *ALKBH8* variants. **(A)** Absence of heterozygosity (AOH) plots for siblings Case 4A (top) and Case 4B (bottom), generated from unphased exome sequencing data using BafCalculator, spanning from base pair 396,909 to 8,102,490 (AOH-chr6:396909-8102490). Both individuals exhibit a 7.7 Mb region of AOH encompassing the *TUBB2B* locus on chromosome 6. Within this region, a red vertical line marks the genomic position chr6:3,226,816 at the variant location. Total autosomal AOH was 435.8 Mb in Case 4A and 324.2 Mb in Case 4B, consistent with parental consanguinity. **(B)** Phenotypic comparison matrix highlighting the contribution of dual molecular diagnoses in the affected siblings with homozygous variants in *TUBB2B* and *ALKBH8*. Red tiles indicate observed phenotypes in each individual. Reference phenotype profiles were compiled from one published case of homozygous *TUBB2B* (blue)^8^, cases of homozygous *ALKBH8* (green)^31^, and heterozygous *TUBB2B* (purple)^7^. Shared features between the two gene associated phenotypes are shown in yellow. This comparative analysis reveals that several neurological and brain imaging features previously attributed to *ALKBH8* (e.g., cerebellar hypoplasia, corpus callosum thinning) are more consistent with the known spectrum of *TUBB2B* associated disease, supporting a dual diagnosis rather than an phenotypic expansion. **(C)** Protein sequence alignment of *TUBB2B* (NM_178012.5) and *TUBB2A* (NM_001069.3) using CLUSTAL O (1.2.4) shows 90% sequence homology. The valine at position 49, indicated by the red arrow, is conserved in both proteins. *Created with BioRender.com*.

The proband’s affected sibling is in the 10-15 year age range, and had global developmental and speech delay, hypotonia, mild hyperactivity, autistic traits, and dysmorphic features (Table 1). Her IQ was 42 (WISC), with limited speech progress despite intervention. Weight and height were more than 2 standard deviations below the mean, and head circumference was between 2 and 3 standard deviations below the mean. She showed no signs of sexual maturation. Brain MRI revealed mild to moderate cerebellar hypoplasia and infratentorial volume loss more pronounced than in the proband, along with ventriculomegaly, corpus callosum thinning, and abnormal myelination.

Neither sibling had a history of seizures, distinguishing them from some previously reported *ALKBH8* cases. Head circumference at birth was not available to determine if there was a primary or secondary microcephaly.

Prior ES identified a homozygous frameshift deletion in *ALKBH8* [Chr11:107375703CG>C (hg19); NM_138775.3: c.1675delC, p.R559Afs*56] predicted to result in a stopgain in the last exon in both affected siblings. Reanalysis revealed an additional novel homozygous missense variant in *TUBB2B* [Chr6:3226816C>T (hg19); NM_178012.5: c.145G>A, p.(V49I)]. This variant was absent from control databases and predicted to be damaging (CADD=25; REVEL=0.52; AlphaMissense=0.57, PrimateAI=0.79) (Table 2), and is in a highly evolutionarily constrained region (Figure 1B).

To assess consanguinity, unphased ES data were analyzed using BafCalculator. Large intervals of AOH were detected: the *TUBB2B* variant lies within a 7.7-Mb block on chromosome 6 in both siblings (Figure 2A), while the previously reported *ALKBH8* variant is located within a 10.6-Mb block in the proband and a 10.8-Mb block in the affected sibling on chromosome 11. The total genome-wide AOH measured 436.4-Mb and 324.7-Mb, respectively. and Sanger sequencing confirmed variant segregation: homozygosity in the affected siblings, and heterozygous in both unaffected parents (Figure 1A).

## Discussion

In this study, we report five individuals from four unrelated families with pathogenic *TUBB2B* variants, each contributing to an expanded allelic and phenotypic spectrum associated with *TUBB2B*-related tubulinopathies. MT stability and plasticity are crucial for neuronal structure and function.

The *TUBB2B* variant in Case 1 is predicted to disrupt tubulin folding and was previously reported in two unrelated individuals in heterozygous form. One, a 10-year-old female with a *de novo* variant, had a complex neurodevelopmental profile including microcephaly, optic atrophy, seizures, scoliosis, polymicrogyria-like cortex, psychomotor delay, corpus callosum agenesis, ventriculomegaly, basal ganglia dysplasia, and mild cereblar and brainstem hypoplasia^23^. The second report describes a fetus with severe lissencephaly characterized by agyria, corpus callosum agenesis, and a normal cerebellum^24^. Our patient shows phenotypic overlap with the previously reported cases, including optic nerve hypoplasia, corpus callosum agenesis, seizures, microcephaly, and developmental delay. These three cases demonstrate the phenotypic variability observed in individuals sharing the same *TUBB2B* variant. We also report panhypopituitarism in Case 1, a feature not previously linked to *TUBB2B* tubulinopathies. Although the optic nerve hypoplasia observed in Case 1 was initially attributed to microcephaly, agenesis of the corpus callosum and panhypopituitarism led to the eventual clinical confirmation of SOD, underscoring the challenges associated with early ascertainment of this diagnosis. Given *TUBB2B*’s role in neuronal migration and early development, this may represent a phenotypic expansion of *TUBB2B*-associated conditions. While the primary manifestations of *TUBB2B* mutations are neurodevelopmental and cortical malformations, pituitary hormone deficiencies may arise when hypothalamic or pituitary structures are affected^25^. This case highlights the need to consider central endocrine involvement as part of the broader clinical presentation for *TUBB2B*. We further identified a rare inherited *PAX2* variant, p.(R195H), associated with papillorenal syndrome (PAPRS [MIM:120330]) in Case 1. While the proband and the carrier parent lack renal findings, this may reflect variable expressivity. The carrier parent was noted to have mild optic nerve hypoplasia, but was otherwise clinically unaffected.

In Case 2 we report a *TUBB2B* variant p.(I202T) previously reported in three individuals^26,27^ allowing detailed phenotype comparison. Shared features with our proband include global developmental delay/intellectual disability (3/3), hypotonia (3/3), hypogenesis or thinning of the corpus callosum (3/3), and cerebellar vermis hypoplasia (3/3). Cortical dysgyria or polymicrogyria was noted in 2 of 3 published cases and also observed in our patient, while microcephaly was documented in 2 of 3 previously and confirmed in our proband. Facial dysmorphism was reported in 2 of 3 cases and is present in our proband, including features such as bilateral ptosis, arched eyebrows, and bow-shaped lips. Behavioral abnormalities, such as hyperactivity and mood lability, were described only in our case. One previously reported individual showed a syndromic presentation with confirmed congenital fibrosis of the extraocular muscles (CFEOM) based on MRI and surgical findings, whereas our proband had bilateral ptosis without ophthalmoplegia, suggesting a milder cranial nerve dysinnervation phenotype. This highlights the phenotypic heterogeneity that can be associated with the same recurrent variant in a tubulin gene and underscores the need for systematic, detailed clinical and neuroimaging assessments in these individuals. Such variability may reflect the influence of additional genetic, epigenetic, or environmental modifiers that shape the final phenotypic expression.

At the molecular level, docking prediction analysis by Mancini *et al.* demonstrated that residue Ile202 plays a crucial role in establishing the hydrophobic environment necessary for GTP stabilization within the β-tubulin nucleotide-binding pocket. Although Ile202 does not directly contact the GTP molecule, its branched, non-polar side chain contributes to the hydrophobic core that maintains pocket integrity^26^. Substitution with threonine, a polar amino acid, is predicted to disrupt this environment, reduce GTP binding affinity, and potentially impair microtubule polymerization. These in silico findings highlight a structurally significant effect of the variant and warrant further experimental validation.

Polyamination, a key tubulin modification, supports MT stability during brain development. While its complete role in neurons is not fully understood, disruptions can impair MT dynamics and contribute to developmental and neurodegenerative disorders^9^. The *TUBB2B* variant in Case 3 affects a conserved polyamination site and has not been previously linked to disease. It is classified as a VUS in ClinVar (Variation ID: 426326) and was reported in one individual with overlapping features such as corpus callosum agenesis, ventriculomegaly, speech delay, and mild intellectual disability who also had a likely pathogenic variant in *MED12L*. Given the variability of brain anomalies in *MED12L* cases, dual diagnosis in this case is possible, though *de novo* status of the variant in *TUBB2B* was not confirmed^28^.

Unlike typical *TUBB2B* variants linked to severe cortical malformations, the Case 3 proband shows a milder phenotype with corpus callosum hypoplasia, a feature seen in other *TUBB2B* cases, suggesting this variant may subtly affect MT stability rather than disrupt assembly. Both increased and decreased MT stability can impair neuronal plasticity. Hyperstabilized MTs may preserve axons but hinder synaptic remodeling, contributing to neuropsychiatric symptoms. Even without major brain malformations, disrupted tubulin polyamination may affect disease by reducing microtubule flexibility and neuronal adaptability.

Multilocus Pathogenic Variation (MPV), for which pathogenic variants at distinct loci contribute to blended phenotypes, are identified in approximately 3.2–7.2% of genetically diagnosed cases^29^. As previously reported, more cases of MPV are expected in populations with increased homozygosity due to distributive AOH burden from consanguinity^16^. Such overlap can mimic phenotypic expansion at a single gene^30^. In Case 4 the brain abnormalities likely reflect a blended phenotype from both *TUBB2B* and *ALKBH8* variants, as similar features have not been observed in other *ALKBH8* cases, ranging from normal brain morphology to macrocephaly and holoprosencephaly^31^. Reanalysis of the exome data identified a novel homozygous *TUBB2B* variant in both the proband and the affected sibling, inherited from heterozygous, clinically unaffected parents. *TUBB2B* is associated with cortical dysplasia complex and brain malformations (CDCBM7 [MIM: 610031]). Most *TUBB2B* variants are heterozygous and missense; there is only one case of a homozygous *TUBB2B* p.(R390Q) variant reported in a Turkish family with three individuals with Uner Tan syndrome^8^. Affected members of this family exhibited a highly diminished cerebellum, causing them to walk with their hands due to lack of balance^8^. Interestingly, this family shares several phenotypic features with our case such as global developmental delay, intellectual disability, hyporeflexia, cerebellar hypoplasia, supratentorial cerebellar volume loss, hypoplasia of corpus callosum, etc. Some features previously attributed to phenotypic expansion of *ALKBH8* are now better explained by the *TUBB2B* variants (Figure 2B). Prior studies have shown that individuals carrying likely pathogenic or pathogenic variants in tubulin genes may exhibit subtle neurological abnormalities on MRI, even in the absence of overt clinical symptoms. Although neuroimaging was not performed for the parents in this case, the presence of a heterozygous *TUBB2B* variant raises the possibility of mild or subclinical neurological phenotypes^32^.

To further investigate the variant’s impact, we examined whether similar variants have been reported in other tubulin genes. *TUBB2A*, a neuron-specific beta-tubulin isotype with 90% homology to *TUBB2B*, is also associated with tubulinopathies (Figure 2C). However, *TUBB2B* shows higher expression during developmental stages. Two unrelated patients have been reported with *de novo TUBB2A* variants at the same amino acid position, p.(V49M) and p.(V49G), showing overlapping features with our case including microcephaly (1/2), dysmorphic features (1/2), intellectual disability (2/2), dysgyria (1/2), thinning of the corpus callosum (1/2), and a small cerebellum (1/2). These reports highlight the pathogenic significance of amino acid changes at this position in *TUBB2B*^33^.

The *TUBB2B* p.(V49I) variant has a lower alpha missense score (0.575) compared to the *TUBB2A* p.(V49G) and p.(V49M) variants (0.959 and 0.993). Suggesting a milder functional effect of the *TUBB2B* p.(V49I) variant, given the structurally similar amino acid substitution, consistent with its presence in unaffected heterozygous parents.

We report three cases with *TUBB2B* variants: a *de novo* variant at a known residue, now linked to an expanded phenotype including panhypopituitarism; a *de novo* heterozygous variant at a polyamination site crucial for microtubule stability; and a novel homozygous missense variant in two siblings, representing another example of MPV, resulting in a blended phenotype with overlapping *ALKBH8*- and *TUBB2B*-related features. Given the varied effects of *TUBB2B* variants on MT dynamics and neuronal development, functional studies are needed to clarify their impact on tubulin structure and function.

## Supplemental information

Supplemental Information includes one table.

## Data Availability

All *TUBB2B* variants have been submitted to ClinVar (Accessions ClinVar:SCV006308805; ClinVar:SCV006308749; ClinVar:SCV006308014). For subjects who have provided written informed consent for sharing of their genomic data in controlled access databases, these data will be deposited to AnVIL and/or dbGaP under accession number dbGaP: phs000711.v5.p1 and dbGaP: phs003047.v3.p2. Access to these data may be granted to qualified researchers who meet the criteria for access to confidential human subject data, subject to approval by the appropriate Institutional Review Board and Data Access Committee at Baylor College of Medicine.

## Acknowledgments

This study was supported in part by the GREGoR Consortium under NIH/NHGRI grant U01HG011758, as well as support from the Caroline Wiess Law Award at Baylor College of Medicine and institutional support from Columbia University Irving Medical Center. Exome sequencing was facilitated by resources at the Human Genome Sequencing Center at Baylor College of Medicine. Additional support was provided through the Genetics & Genomics Training Program (T32GM139534) funded by the National Institute of General Medical Sciences. The data for Case 2 were obtained while Dr. Sare Nur Yılmaz was affiliated with Istanbul Medeniyet University. We thank the patients and their families for their participation in this research.

## Declaration of Interests

Jennifer E. Posey serves on the Scientific Advisory Board of MaddieBio. Davut Pehlivan provides consulting service for Ionis Pharmaceuticals, M2DS Therapeutics and Acadia Pharmaceuticals. Divya Kalra’s immediate family member is employed at Oxford Nanopore Technologies. The Department of Molecular and Human Genetics at Baylor College of Medicine receives revenue from clinical genetic testing completed at Baylor Genetics Laboratory. The remaining authors declare that they have no competing interests.

## Web Resources

https://stuart.radboudumc.nl/metadome/

http://www.omim.org/

https://gnomad.broadinstitute.org/

http://lilab2.sysu.edu.cn/Tools/msa/clustalo/

## Notes

### Author Declarations

Ethics committee/IRB of Baylor College of Medicine gave ethical approval for this work

## References

1. Binarová, P., and Tuszynski, J. (2019). Tubulin: structure, functions and roles in disease. Cells 8. 10.3390/cells8101294.

2. Tantry, M.S.A., and Santhakumar, K. (2023). Insights on the Role of α- and β-Tubulin Isotypes in Early Brain Development. Mol. Neurobiol. 60, 3803–3823. 10.1007/s12035-023-03302-1.

3. Poretti, A., Boltshauser, E., and Huisman, T.A.G.M. (2014). Congenital brain abnormalities: an update on malformations of cortical development and infratentorial malformations. Semin. Neurol. 34, 239–248. 10.1055/s-0034-1386762.

4. Mitani, T., Punetha, J., Akalin, I., Pehlivan, D., Dawidziuk, M., Coban Akdemir, Z., Yilmaz, S., Aslan, E., Hunter, J.V., Hijazi, H., et al. (2019). Bi-allelic Pathogenic Variants in *TUBGCP2* Cause Microcephaly and Lissencephaly Spectrum Disorders. Am. J. Hum. Genet. 105, 1005–1015. 10.1016/j.ajhg.2019.09.017.

5. Poirier, K., Saillour, Y., Bahi-Buisson, N., Jaglin, X.H., Fallet-Bianco, C., Nabbout, R., Castelnau-Ptakhine, L., Roubertie, A., Attie-Bitach, T., Desguerre, I., et al. (2010). Mutations in the neuronal ß-tubulin subunit *TUBB3* result in malformation of cortical development and neuronal migration defects. Hum. Mol. Genet. 19, 4462–4473. 10.1093/hmg/ddq377.

6. Tischfield, M.A., Baris, H.N., Wu, C., Rudolph, G., Van Maldergem, L., He, W., Chan, W.-M., Andrews, C., Demer, J.L., Robertson, R.L., et al. (2010). Human *TUBB3* mutations perturb microtubule dynamics, kinesin interactions, and axon guidance. Cell 140, 74–87. 10.1016/j.cell.2009.12.011.

7. Tischfield, M.A., Cederquist, G.Y., Gupta, M.L., and Engle, E.C. (2011). Phenotypic spectrum of the tubulin-related disorders and functional implications of disease-causing mutations. Curr. Opin. Genet. Dev. 21, 286–294. 10.1016/j.gde.2011.01.003.

8. Breuss, M.W., Nguyen, T., Srivatsan, A., Leca, I., Tian, G., Fritz, T., Hansen, A.H., Musaev, D., McEvoy-Venneri, J., James, K.N., et al. (2017). Uner Tan syndrome caused by a homozygous *TUBB2B* mutation affecting microtubule stability. Hum. Mol. Genet. 26, 258–269. 10.1093/hmg/ddw383.

9. Janke, C. (2014). The tubulin code: molecular components, readout mechanisms, and functions. J. Cell Biol. 206, 461–472. 10.1083/jcb.201406055.

10. Saad, A.K., Marafi, D., Mitani, T., Du, H., Rafat, K., Fatih, J.M., Jhangiani, S.N., Coban-Akdemir, Z., Baylor-Hopkins Center for Mendelian Genomics, Gibbs, R.A., et al. (2021). Neurodevelopmental disorder in an Egyptian family with a biallelic ALKBH8 variant. Am. J. Med. Genet. A 185, 1288–1293. 10.1002/ajmg.a.62100.

11. Posey, J.E., O’Donnell-Luria, A.H., Chong, J.X., Harel, T., Jhangiani, S.N., Coban Akdemir, Z.H., Buyske, S., Pehlivan, D., Carvalho, C.M.B., Baxter, S., et al. (2019). Insights into genetics, human biology and disease gleaned from family based genomic studies. Genet. Med. 21, 798–812. 10.1038/s41436-018-0408-7.

12. Bayram, Y., Gulsuner, S., Guran, T., Abaci, A., Yesil, G., Gulsuner, H.U., Atay, Z., Pierce, S.B., Gambin, T., Lee, M., et al. (2015). Homozygous loss-of-function mutations in *SOHLH*1 in patients with nonsyndromic hypergonadotropic hypogonadism. J. Clin. Endocrinol. Metab. 100, E808–14. 10.1210/jc.2015-1150.

13. Reid, J.G., Carroll, A., Veeraraghavan, N., Dahdouli, M., Sundquist, A., English, A., Bainbridge, M., White, S., Salerno, W., Buhay, C., et al. (2014). Launching genomics into the cloud: deployment of Mercury, a next generation sequence analysis pipeline. BMC Bioinformatics 15, 30. 10.1186/1471-2105-15-30.

14. Farek, J., Hughes, D., Salerno, W., Zhu, Y., Pisupati, A., Mansfield, A., Krasheninina, O., English, A.C., Metcalf, G., Boerwinkle, E., et al. (2022). xAtlas: scalable small variant calling across heterogeneous next-generation sequencing experiments. Gigascience 12. 10.1093/gigascience/giac125.

15. Pehlivan, D., Bayram, Y., Gunes, N., Coban Akdemir, Z., Shukla, A., Bierhals, T., Tabakci, B., Sahin, Y., Gezdirici, A., Fatih, J.M., et al. (2019). The genomics of arthrogryposis, a complex trait: candidate genes and further evidence for oligogenic inheritance. Am. J. Hum. Genet. 105, 132–150. 10.1016/j.ajhg.2019.05.015.

16. Coban-Akdemir, Z., Song, X., Ceballos, F.C., Pehlivan, D., Karaca, E., Bayram, Y., Mitani, T., Gambin, T., Bozkurt-Yozgatli, T., Jhangiani, S.N., et al. (2024). The impact of the Turkish population variome on the genomic architecture of rare disease traits. Genetics in Medicine Open 2, 101830. 10.1016/j.gimo.2024.101830.

17. Olshen, A.B., Venkatraman, E.S., Lucito, R., and Wigler, M. (2004). Circular binary segmentation for the analysis of array-based DNA copy number data. Biostatistics 5, 557–572. 10.1093/biostatistics/kxh008.

18. Rentzsch, P., Witten, D., Cooper, G.M., Shendure, J., and Kircher, M. (2019). CADD: predicting the deleteriousness of variants throughout the human genome. Nucleic Acids Res. 47, D886–D894. 10.1093/nar/gky1016.

19. Minton, K. (2023). Predicting variant pathogenicity with AlphaMissense. Nat. Rev. Genet. 24, 804. 10.1038/s41576-023-00668-9.

20. Ioannidis, N.M., Rothstein, J.H., Pejaver, V., Middha, S., McDonnell, S.K., Baheti, S., Musolf, A., Li, Q., Holzinger, E., Karyadi, D., et al. (2016). REVEL: an ensemble method for predicting the pathogenicity of rare missense variants. Am. J. Hum. Genet. 99, 877–885. 10.1016/j.ajhg.2016.08.016.

21. Fromer, M., and Purcell, S.M. (2014). Using XHMM Software to Detect Copy Number Variation in Whole-Exome Sequencing Data. Curr. Protoc. Hum. Genet. 81, 7.23.1-7.23.21. 10.1002/0471142905.hg0723s81.

22. Gambin, T., Akdemir, Z.C., Yuan, B., Gu, S., Chiang, T., Carvalho, C.M.B., Shaw, C., Jhangiani, S., Boone, P.M., Eldomery, M.K., et al. (2017). Homozygous and hemizygous CNV detection from exome sequencing data in a Mendelian disease cohort. Nucleic Acids Res. 45, 1633–1648. 10.1093/nar/gkw1237.

23. Cushion, T.D., Dobyns, W.B., Mullins, J.G.L., Stoodley, N., Chung, S.-K., Fry, A.E., Hehr, U., Gunny, R., Aylsworth, A.S., Prabhakar, P., et al. (2013). Overlapping cortical malformations and mutations in *TUBB2B* and *TUBA1A*. Brain 136, 536–548. 10.1093/brain/aws338.

24. Fallet-Bianco, C., Laquerrière, A., Poirier, K., Razavi, F., Guimiot, F., Dias, P., Loeuillet, L., Lascelles, K., Beldjord, C., Carion, N., et al. (2014). Mutations in tubulin genes are frequent causes of various foetal malformations of cortical development including microlissencephaly. Acta Neuropathol. Commun. 2, 69. 10.1186/2051-5960-2-69.

25. Oren, N.C., Cag ltay, E., Akay, F., and Toyran, S. (2015). Panhypopituitarism with Ectopic Posterior Pituitary Lobe, Heterotopia, Polymicrogyria, Corpus Callosum Dysgenesis, and Optic Chiasm/Nerve Hypoplasia: Is That an Undefined Neuronal Migration Syndrome? AJNR Am J Neuroradiol 36, E33–E35. 10.3174/ajnr.A4305.

26. Mancini, C., Chiriatti, L., Bruselles, A., D’ambrosio, P., Ciolfi, A., Ferilli, M., Cappelletti, C., Carvetta, M., Radio, F.C., Cordeddu, V., et al. (2025). The p.Ile202Thr Substitution in *TUBB2B* Can Be Associated with Syndromic Presentation of Congenital Fibrosis of the Extraocular Muscles. Genes 16. 10.3390/genes16101182.

27. Bah i-Buisson, N., Poirier, K., Fourniol, F., Saillour, Y., Valence, S., Lebrun, N., Hully, M., Bianco, C.F., Boddaert, N., Elie, C., et al. (2014). The wide spectrum of tubulinopathies: what are the key features for the diagnosis? Brain 137, 1676–1700. 10.1093/brain/awu082.

28. Nizon, M., Laugel, V., Flanigan, K.M., Pastore, M., Waldrop, M.A., Rosenfeld, J.A., Marom, R., Xiao, R., Gerard, A., Pichon, O., et al. (2019). Variants in *MED12L*, encoding a subunit of the mediator kinase module, are responsible for intellectual disability associated with transcriptional defect. Genet. Med. 21, 2713–2722. 10.1038/s41436-019-0557-3.

29. Mitani, T., Isikay, S., Gezdirici, A., Gulec, E.Y., Punetha, J., Fatih, J.M., Herman, I., Akay, G., Du, H., Calame, D.G., et al. (2021). High prevalence of multilocus pathogenic variation in neurodevelopmental disorders in the Turkish population. Am. J. Hum. Genet. 108, 1981–2005. 10.1016/j.ajhg.2021.08.009.

30. Karaca, E., Posey, J.E., Coban Akdemir, Z., Pehlivan, D., Harel, T., Jhangiani, S.N., Bayram, Y., Song, X., Bahrambeigi, V., Yuregir, O.O., et al. (2018). Phenotypic expansion illuminates multilocus pathogenic variation. Genet. Med. 20, 1528–1537. 10.1038/gim.2018.33.

31. Waqas, A., Nayab, A., Shaheen, S., Abbas, S., Latif, M., Rafeeq, M.M., Al-Dhuayan, I.S., Alqosaibi, A.I., Alnamshan, M.M., Sain, Z.M., et al. (2022). Case Report: Biallelic Variant in the tRNA Methyltransferase Domain of the AlkB Homolog 8 Causes Syndromic Intellectual Disability. Front. Genet. 13, 878274. 10.3389/fgene.2022.878274.

32. Durizot, M., Burglen, L., Garel, C., Blondiaux, E., Riquet, A., Floret, V., Desportes, V., Häänpaa, M., Valenzuela, M.I., Pinto, A.M., et al. (2025). Attenuated clinical forms of tubulinopathies in children and adults: A series of 24 individuals. Pediatr. Neurol. 170, 49–57. 10.1016/j.pediatrneurol.2025.06.003.

33. Brock, S., Vanderhasselt, T., Vermaning, S., Keymolen, K., Régal, L., Romaniello, R., Wieczorek, D., Storm, T.M., Schaeferhoff, K., Hehr, U., et al. (2021). Defining the phenotypical spectrum associated with variants in *TUBB2A*. J. Med. Genet. 58, 33–40. 10.1136/jmedgenet-2019-106740.

